# Receipt of guideline-concordant prenatal care among Medicaid-insured women with disabilities in Texas

**DOI:** 10.64898/2026.10.25.26366331

**Authors:** Alka Dev, Andrew Schaefer, Audrey Douches, Willi Horner-Johnson, Thérèse A Stukel, David C. Goodman, JoAnna K. Leyenaar

## Abstract

**Objective:** To assess receipt of guideline-concordant prenatal care among pregnant women with disabilities enrolled in Medicaid.

**Methods:** We conducted a retrospective cohort study of 748,304 births to women enrolled in Texas Medicaid from 2010–2015. Births were classified into three groups by maternal disability status: no disability, receipt of federal disability benefits (STAR+), or disability identified through ICD-9 codes. Guideline-concordant care was defined as receipt of recommended prenatal services at recommended times. Multivariable logistic regression estimated adjusted odds ratios (aORs) for guideline-concordant care, adjusting for covariates.

**Results:** There were 20,700 births (2.8%) to women with disabilities, 40.1% of whom were enrolled in STAR+. Fewer than half of women in any group received guideline-concordant testing for D(Rh) antigen, rubella, complete blood count, bacteriuria, or glucose tolerance. Women in STAR+ were more likely than those without disabilities to receive no prenatal care (9.4% vs. 1.6%), despite higher first-trimester initiation among those who accessed care. STAR+ enrollment was associated with substantially lower odds of guideline-concordant testing, including HIV (aOR 0.27; 95% CI, 0.26–0.29), syphilis (0.33; 0.31–0.35), hepatitis B (0.38; 0.36–0.40), D(Rh) antigen (0.31; 0.30–0.33), anatomy ultrasound (0.32; 0.31–0.34), and glucose (0.42; 0.40–0.44). Women with disabilities identified via ICD-9 codes also had lower odds, though differences were smaller.

**Conclusion for practice:** Guideline-concordant prenatal care was suboptimal overall and especially deficient among women receiving federal disability benefits, underscoring the need for targeted quality improvement in Medicaid prenatal care delivery.

**Significance:** *What is already known on this subject?:* Pregnant women with disabilities face substantially higher risks of severe maternal morbidity, gestational complications, and adverse infant outcomes. Evaluating whether Medicaid delivers guideline-concordant prenatal care to this population is essential for identifying care gaps in a major safety-net program.

*What this study adds:* In this cohort study, receipt of guideline-concordant prenatal care was low overall and significantly lower among women with disabilities. Women receiving federal disability benefits (STAR+) had the lowest concordance rates and were more likely to receive no prenatal care compared to women without disabilities.

## INTRODUCTION

Pregnant women with disabilities experience significantly higher rates of complications such as severe maternal morbidity, gestational diabetes, preeclampsia, and postpartum hemorrhage, regardless of disability type ^1–3^. Their infants are similarly at increased risk, with higher rates of being small for gestational age, premature, or low birthweight ^4–6^. These disparities may stem, in part, from different patterns of healthcare utilization during pregnancy, with some evidence of delayed initiation of prenatal care and fewer total prenatal care visits in this population when compared to women without disabilities ^7,8^. Having a disability may expose women to unique structural, systemic, and interpersonal barriers that impede access to adequate prenatal care. As a result, care is often neither timely nor sufficient, and may be irresponsive to the specific needs of individuals with disabilities ^9–11^. Communication challenges between patients and providers, combined with healthcare facilities that lack adequate accommodations, may further limit the quality and continuity of care received throughout the perinatal period ^12,13^. Collectively, these disparities underscore a critical need to identify opportunities to improve maternal healthcare delivery for pregnant women with disabilities.

Quantifying prenatal care adequacy through measures of timing and frequency provides only a partial assessment of care quality ^14^. Guideline-concordant prenatal care, which assesses whether recommended specific clinical services are delivered at the appropriate gestational age, is an enhanced measure of prenatal care quality ^15^. Understanding health service utilization during pregnancy provides insight into healthcare access and quality, particularly for populations at heightened risk of adverse maternal and neonatal outcomes ^16,17^. Such analyses are especially relevant for evaluating the performance of critical safety-net programs like Medicaid, which provides coverage for approximately one-third of Americans with disabilities and finances more than 40% of births nationally ^18,19^. In this study, we aimed to determine whether pregnant women with disabilities enrolled in Medicaid received guideline-concordant prenatal care. We examined utilization patterns across three groups: pregnant women with disabilities who received federal disability benefits, those with disabilities who were not receiving disability benefits, and those with no documented disabilities. Given the date of our data, this serves as a baseline assessment to identify care gaps for monitoring prenatal care trends in this population.

## METHODS

### Study Design and Data Source

We conducted a retrospective cohort study of livebirths to women enrolled in Texas Medicaid from 2010 to 2015. The cohort was derived from linked maternal and infant Medicaid claims, enrollment files, and birth certificates. Maternal files included Medicaid and disability eligibility and facility and professional claims for up to 12 months before delivery, the delivery hospitalization, and two months postpartum. Birth certificates provided maternal age, education, race, ethnicity, marital status, and nativity.

The study received approval from the institutional review boards of Dartmouth College, with a waiver of informed consent according to 45 CFR 46.116(d). The study adhered to the Strengthening the Reporting of Observational Studies in Epidemiology (STROBE) reporting guidelines for cohort studies ^**20**^.

### Population

We included 748,304 singleton livebirths to women aged 15–44 years delivered between 22 and 44 weeks’ gestation (Figure 1). We excluded births with missing maternal linkage (n=100,606), fewer than 3 months of continuous enrollment including the birth month (n=376,339), no maternal claims (n=5,848), or no ambulatory encounters during pregnancy (n=10,840). Given the large number of exclusions due to a lack of 3 months of continuous enrollment, we also described the demographic characteristics of this excluded group (Appendix 1).

**Figure 1.**
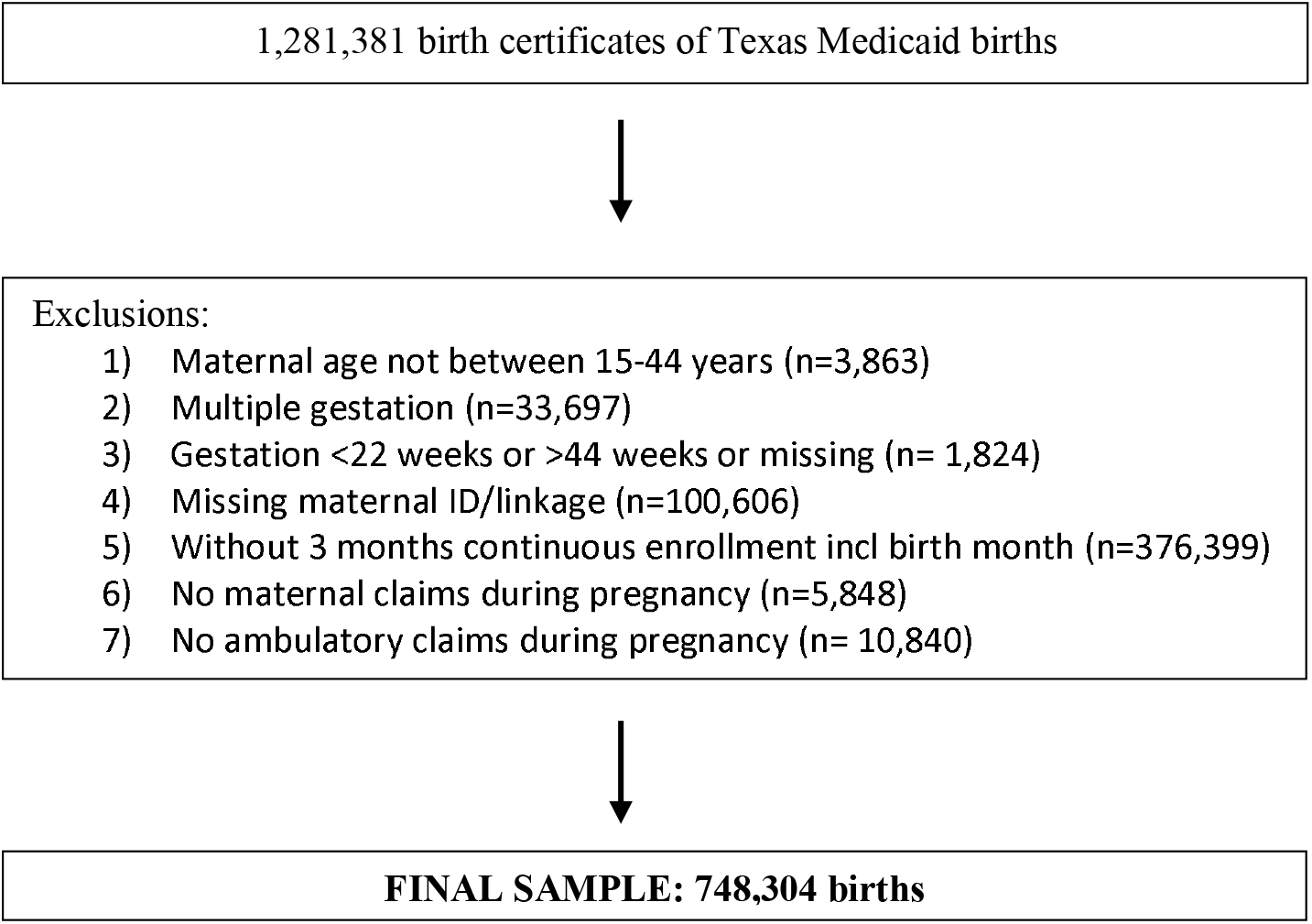
Sample Selection.

### Disability identification and Medicaid eligibility

In Texas, women qualifying for Supplemental Security Income (SSI) receive pregnancy coverage through the STAR+ Medicaid program.^21^ We previously described how we identified maternal disability based on either enrollment in STAR+ or having at least one disability associated with an International Classification of Diseases – Ninth Edition (ICD-9) code recorded during two ambulatory visits during pregnancy, one emergency room visit, or one hospitalization ^22,23^. For the current analysis, we created three groups for comparison: births to women with no documented disabilities, births to women enrolled in STAR+ (federal disability benefits), and births to women with only ICD-9 codes for disability conditions. Women with both STAR+ enrollment and ICD-9 codes for disability were included in the STAR+ group.

### Ambulatory care utilization

The pregnancy period was established based on the estimated date of the last menstrual period (calculated as ‘delivery date minus gestation in weeks’ from birth certificate files). Ambulatory care included all outpatient visits during the pregnancy period and was identified using “place of service” codes from professional claims and “type of bill” codes from facility claims (Appendix 2). These included visits to physician offices, federally qualified health centers, rural health clinics, urgent care facilities, telehealth, and critical care hospitals.

### Prenatal care utilization

Claims data can measure perinatal health care receipt, content, timing, and quality in greater detail than traditional vital statistics measures, if prenatal care and delivery are not paid as a bundle ^24^. We measured prenatal care timing, visit counts, and guideline-concordant services. Prenatal care visits were identified from ambulatory visits using three ICD-9 codes: ‘Normal pregnancy’ (v22.0, v22.1, v22.2); ‘Supervision of high risk pregnancy’ (v23.1-v23.5, v23.7-v23.9); and ‘Antenatal screening’ (v28.0-v28.6, v28.8-v28.9). The timing of prenatal care initiation was measured as the trimester in which the individual had their first prenatal care visit. The number of visits was the total count of unique visits with a physician, certified nurse midwife or midwife, nurse practitioner, or physician’s assistant.

Guideline-concordant prenatal care was measured using procedures and services delivered at designated prenatal care visits as described by Gourevitch and colleagues based on recommendations set by the American College of Obstetricians and Gynecologists, the Centers for Disease Control, and Prevention, the US Preventive Services Task Force, and/or the International Society of Ultrasound in Obstetrics and Gynecology (Appendix 2) ^15^. We counted visits to independent labs as separate from the most recent provider visit if more than one week elapsed, accommodating potential billing delays.

Receipt of prenatal care was considered ‘on time’ if received as recommended, and ‘mistimed’ if a patient was enrolled in Medicaid at the recommended gestation but received the test at some other time. We also report data for patients who did not receive the test at all (designated ‘not done’). Finally, we summarized the types of providers seen during prenatal care appointments.

For those with facility only prenatal care claims, the type of provider was generally listed as the facility type, i.e. hospital, primary care facility, or ambulatory care facility (designated ‘other’ in our analysis).

### Covariates

Maternal sociodemographic characteristics, self-reported on the birth certificate, included age, education, race, ethnicity, and marital status. Age was reported as a linear variable as well as 10 year categories between 15-44 years. Education categories included less than high school, high school, and some college or higher. Race categories included Asian, Black, White, more than one race, and Other, which included American Indian, Pacific Islanders, Other, and Unknown race. Hispanic ethnicity was measured separately; thus, women could identify as Hispanic and designate a race. Marital status referred to being married or not at the time of birth. We also examined smoking during pregnancy, given the differences in smoking rates among pregnant people with and without disabilities ^4,25^.

### Statistical Analysis

We stratified all descriptive analyses by the two disability groups and the no disability group as the reference population. Adjusted odds ratios (aORs) and 95% confidence intervals (CIs) for receipt of guideline-concordant care (yes/no) were estimated using multivariable logistic regression, adjusting for the number of Medicaid enrollment months during pregnancy (possible range 1-10), age in years, and categories for race, ethnicity, rurality, and education. We used the beneficiary ID to account for the clustering of pregnancies within women ^26^. Analyses were performed using the PROC GENMOD using SAS version 9.4 (SAS Institute, Cary, NC) during April – November 2025.

## RESULTS

### Sample Characteristics

The final sample included 748,304 singleton livebirths to 622,499 women (Table 1). Of these, 20,700 births (2.8%) were to women with disabilities identified by STAR+ or ICD-9 codes. The majority of women with disabilities (n=12,396, 59.9%) were identified through ICD-9 codes only, while 8,304 (40.1%) were enrolled in STAR+; approximately 0.22% (1,609) of STAR+ members also had ICD-9 codes. Physical disabilities were most common (55.5% of women with disabilities), followed by sensory (9.6%) and intellectual disabilities (5.2%). Notably, 29.7% of women with disabilities had no documented disability ICD-9 codes but were enrolled in STAR+.

**Table 1:** Sample characteristics by disability group.

|  | No disability | All STAR+<br>(incl ICD-9) | ICD-9 Only | TOTAL |
| --- | --- | --- | --- | --- |
| No. of births | 727,604<br>(97.2%) | 8,304<br>(1.1%) | 12,396<br>(1.7%) | 748,304 |
| No. of women<sup>a</sup> | 606,644 | 7,221 | 12,091 | 622,499 |
| Mean enrollment months during pregnancy (SD) | 7.7 (1.7) | 9.2 (0.8) | 8.2 (1.5) | 7.7 (1.7) |
| **Type of disability** |  |  |  |  |
| Physical | -- | 1,259 (15.2%) | 10,223<br>(82.5%) | 11,482 (1.5%) |
| Intellectual | -- | 191 (2.3%) | 875 (7.1%) | 1,066 (0.1%) |
| Sensory | -- | 315 (3.8%) | 1,681 (13.6%) | 1,996 (0.3%) |
| Not documented | 727,604 (100%) | 6,539 (78.7%) | -- | 733,760 (98.1%) |
| **Age** |  |  |  |  |
| Mean (SD) | 24.5 (5.5) | 25.4 (5.6) | 25.3 (5.9) | 24.6 (5.5) |
| 15-24 years | 411,969 (56.6%) | 4,195 (50.5%) | 6,261 (50.5%) | 422,425 (56.5%) |
| 25-34 years | 272,958 (37.5%) | 3,462 (41.7%) | 5,096 (41.1%) | 281,516 (37.6%) |
| 35-44 years | 42,677 (5.9%) | 647 (7.8%) | 1,039 (8.4%) | 44,363 (5.9%) |
| **Education<sup>b</sup>** |  |  |  |  |
| < High school | 182,054 (25.0%) | 3,448 (41.6%) | 3,158 (25.5%) | 188,660 (25.2%) |
| Completed high school | 278,218 (38.3%) | 3,201 (38.6%) | 4,701 (38.0%) | 286,120 (38.3%) |
| ≥ Some college | 266,772 (36.7%) | 1,642 (19.8%) | 4,519 (36.5%) | 272,933 (36.5%) |
| **Race** |  |  |  |  |
| Asian | 12,161 (1.7%) | 71 (0.9%) | 156 (1.3%) | 12,388 (1.7%) |
| Black | 124,224 (17.1%) | 2,770 (33.4%) | 2,161 (17.4%) | 129,155 (17.3%) |
| White | 489,375 (67.3%) | 4,409 (53.1%) | 8,648 (69.8%) | 502,432 (67.1%) |
| More than one race | 10,496 (1.4%) | 142 (1.7%) | 228 (1.9%) | 10,866 (1.5%) |
| All Other / Unknown | 91,348 (12.6%) | 912 (11.0%) | 1,203 (9.7%) | 93,463 (12.5%) |
| **Ethnicity** |  |  |  |  |
| Hispanic | 382,899 (52.6%) | 3,761 (45.3%) | 5,130 (41.4%) | 391,790 (52.4%) |
| **Parity<sup>b</sup>** |  |  |  |  |
| 1 | 287,180 (39.5%) | 2,895 (34.9%) | 5,230 (42.2%) | 295,305 (39.5%) |
| 2 | 208,600 (28.7%) | 2,140 (25.8%) | 3,323 (26.8%) | 214,063 (28.6%) |
| ≥ 3 | 231,642 (31.8%) | 3,264 (39.3%) | 3,838 (31.0%) | 238,744 (31.9%) |
| **Married** | 257,021 (35.3%) | 2,165 (26.1%) | 4,219 (34.0%) | 263,405 (35.2%) |
| **Smoking during pregnancy (%)** | 58,203 (8.0%) | 812 (9.8%) | 1,499 (12.1%) | 60,514 (8.1%) |
| **Rural Residence (%)** | 111,939 (15.4%) | 573 (6.9%) | 1,983 (16.0%) | 114,495 (15.3%) |
| **Mental Illness** | 26,053 (3.6%) | 2,416 (29.1%) | 2,390 (19.3%) | 30,859 (4.1%) |

<sup>a</sup> Women can be in >1 group if they have multiple births in the sample; cells will not add up to the total.
<sup>b</sup> 591 were missing education; 192 were missing parity.

Compared to those without disabilities, educational attainment was lowest among those in the STAR+ group, with 41.6% having less than a high school education compared to 25.0% of women without disabilities, and only 19.8% having some college education versus 36.7% in the non-disabled group. Racial composition varied, with a higher proportion of Black women in the STAR+ group (33.4% compared with 17.1% of those without disabilities). Rural residence was observed most infrequently in the STAR+ group (6.9%) compared to the no disability group (15.4%). Mental illness prevalence was substantially higher among women in STAR+ (29.1% vs. 3.6% for no disability).

### Health Services Utilization

Women in STAR+ averaged 18.0 total ambulatory visits during pregnancy, those in the ICD-9 disability group averaged 21.0 visits, and those with no disabilities averaged 14.5 visits (Table 2). The mean number of prenatal care visits was lowest in the STAR+ group (10.9) and highest in the ICD-9 group (13.5), compared to the no disability group (12.2). The timing of prenatal care initiation differed between groups as well. A greater proportion of women in the STAR+ and ICD-9 groups initiated care in the first trimester (72.1% and 72.2% respectively vs. 64.3% for no disability). However, women in the STAR+ group had a notably higher rate of never receiving prenatal care (9.4% vs. 1.6% for no disability). Among women with ICD-9 codes only, 4.4% never received prenatal care. The distribution of prenatal care provider types was similar across all three groups.

**Table 2:** Health services utilization by disability group.

|  | No disability | All STAR+<br>(incl ICD-9) | ICD-9 Only | TOTAL |
| --- | --- | --- | --- | --- |
| **Outpatient healthcare visits during pregnancy** |  |  |  |  |
| Ambulatory, mean (SD) | 14.5 (7.4) | 18.0 (15.2) | 21.0 (15.4) | 14.6 (7.8) |
| Prenatal, mean (SD) | 12.2 (5.8) | 10.9 (8.0) | 13.5 (7.8) | 12.2 (5.8) |
| **Timing of prenatal care initiation** |  |  |  |  |
| First trimester | 467,630<br>(64.3%) | 5,990<br>(72.1%) | 8,953<br>(72.2%) | 482,573<br>(64.5%) |
| Second trimester | 207,934<br>(28.6%) | 1,202<br>(14.5%) | 2,474<br>(20.0%) | 211,610<br>(28.3%) |
| Third trimester | 40,342 (5.5%) | 332 (4.0%) | 427 (3.4%) | 41,101 (5.5%) |
| Never (no prenatal care codes) | 11,698 (1.6%) | 780 (9.4%) | 542 (4.4%) | 13,020 (1.7%) |
| **Type of PNC provider (of all PNC visits from claims) %** |  |  |  |  |
| Physician | 5,884,313<br>(66.4%) | 59,053<br>(64.5%) | 107,884<br>(64.3%) | 6,051,250<br>(64.3%) |
| Nurse Practitioner/Physician Assistant | 360,286<br>(4.1%) | 3,050<br>(3.3%) | 6,376<br>(1.7%) | 369,712<br>(3.8%) |
| Certified Midwife/Nurse Midwife | 13,702<br>(0.2%) | 60<br>(0.4%) | 146<br>(1.1%) | 13,908<br>(0.2%) |
| Other | 2,603,923<br>(29.4%) | 29,441<br>(31.1%) | 53,424<br>(31.8%) | 2,686,788<br>(29.5%) |

### Prenatal Care Concordance

Figure 2 presents care concordance rates across recommended services (table in Appendix 4). Tests recommended at any time during pregnancy were, by definition, concordant if a woman received the test at all; these included HIV, syphilis, and hepatitis B testing. Concordance rates for these tests were lowest for those in the STAR+ group (69.4 - 71.5%) and ranged from 72.6 - 84.7% for those with ICD-9 disability codes or no disability. Fewer than 50% of any pregnant women received screening tests for D(Rh) antigen, red blood cell count (RBC), rubella, complete blood count (CBC), bacteriuria, or oral glucose tolerance, but rates were especially low among STAR+ beneficiaries, ranging from 16.9% for rubella screening to 34.8% for bacteriuria. Receipt of the TDAP vaccine was the lowest of any test or intervention across all groups, with less than 10% of all women receiving the vaccine on time. Anatomy scans had the lowest concordance for women in STAR+ (47.5%) compared to those with ICD-9 codes (63.2%) and women without disabilities (64.6%). Similarly, only 36.8% of women in STAR+ half of women received Group B Streptococcus testing as recommended compared to those with ICD-9 codes (52.0%) or without disability (58.4%).

**Figure 2:**
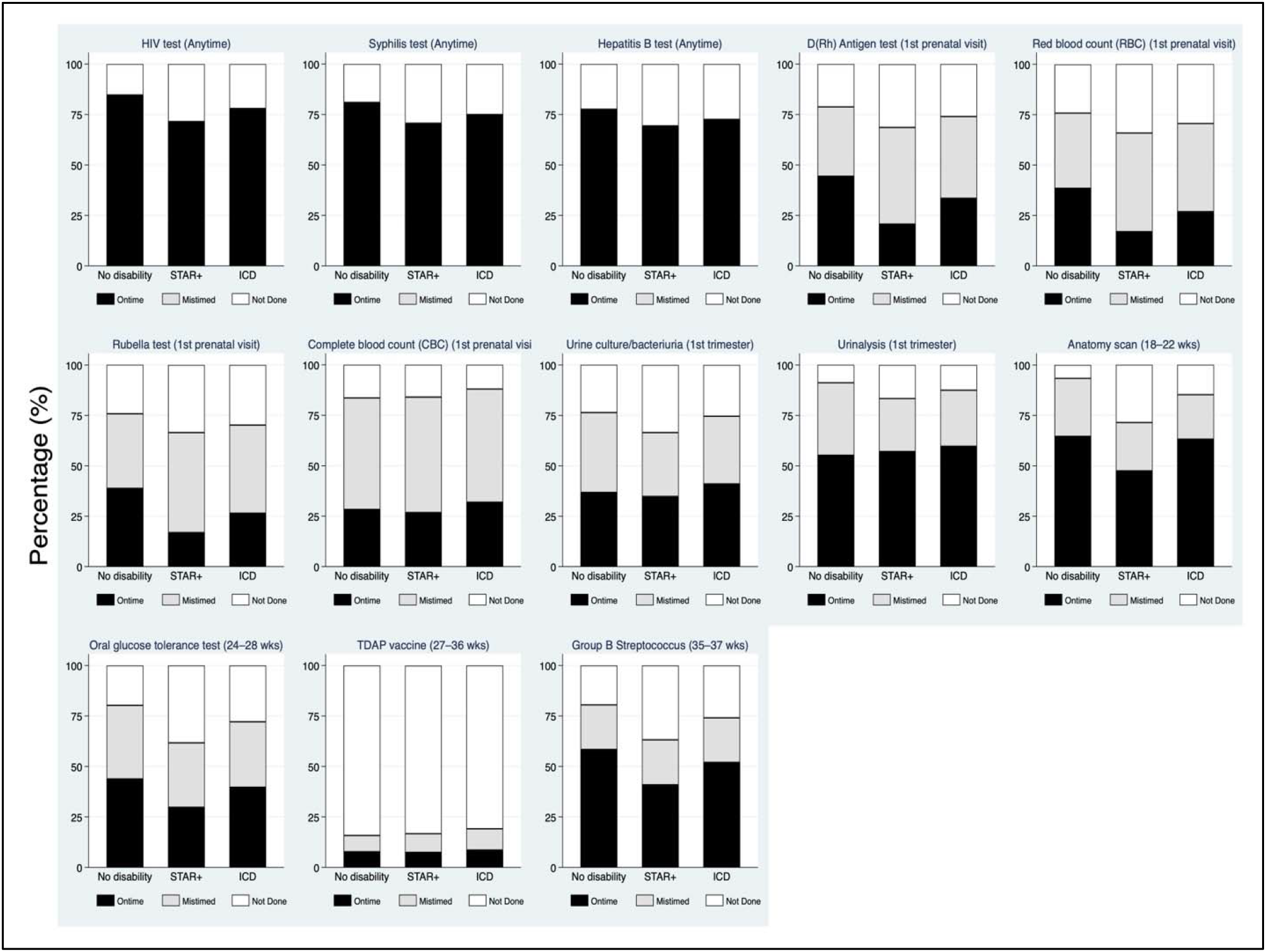
Concordance of tests during recommended gestation windows.

Overall, women enrolled in STAR+ had lower odds of receiving guideline concordant care compared to women with no disability. STAR+ enrollment was associated with lower odds of screening for HIV (aOR 0.27; 95% CI, 0.26–0.29), syphilis (0.33; 0.31–0.35), and hepatitis B (0.38; 0.36–0.40). Similarly lower odds were observed among STAR+ women for first-trimester laboratory services, including D(Rh) antigen testing (0.31; 0.30–0.33), RBC (0.33; 0.31–0.35), rubella screening (0.33; 0.31–0.35), as well as anatomy ultrasound at 18–22 weeks (0.32; 0.31– 0.34) and glucose testing at 24–28 weeks (0.42; 0.40–0.44).

In the ICD-9 disability group, odds of receiving HIV (0.55, 95% CI= 0.52–0.57), syphilis (0.59, 95% CI= 0.57–0.62), and hepatitis B (0.65, 95% CI= 0.62–0.67) testing were lower compared to those without disability, though differences were smaller in magnitude than those observed in the STAR+ group. A similar pattern was apparent for most other services, including D(Rh) antigen testing (0.62; 0.60–0.65), RBC (0.59; 0.56–0.61), and rubella screening (0.57; 0.55–0.59). Anatomy ultrasound (0.84; 0.80–0.87) and glucose screening (0.78; 0.75–0.81) were also less likely compared with those without disability.

## DISCUSSION

This study found significant gaps in adherence to evidence-based prenatal care guidelines among Medicaid insured pregnant women, with receipt of guideline-concordant care significantly lower among those with disabilities compared to women without disabilities. However, guideline-concordant care was low across all groups with few exceptions, indicating that Medicaid-enrolled pregnant women receive low quality care in general, with particularly low quality for women with disabilities. A higher proportion of women in STAR+ and those with ICD-9 disability codes started prenatal care in the first trimester compared to women without reported disabilities. This may reflect greater healthcare system contact, as shown by their higher number of ambulatory care visits.

Overall, initiation of prenatal care in the first trimester was generally high in the U.S. during this period. Data from the 2016 Pregnancy Risk Assessment Monitoring System (PRAMS) showed that, nationally, 92% of U.S. women initiated prenatal care in the first trimester in 2016, but only 78.6% of Medicaid beneficiaries did so, compared to 93.8% with private insurance.^27^ Our finding that less than two thirds of women enrolled in Texas Medicaid during this time received timely prenatal care, regardless of disability status, reflects below-average care quality. These rates are also much lower than those found by Gourevitch and colleagues in their analysis using commercial insurance data.

Women enrolled in STAR+ were more likely to receive no prenatal care at all than women without disabilities and those with ICD-9 disability codes. Previous research from Ontario showed that inadequate prenatal care increased with disability severity despite higher overall care intensity ^8^. In alignment with that finding, our results suggest two groups: women with disabilities who successfully initiate prenatal care and a smaller group that faces barriers to accessing any prenatal care. Research from California found that women with intellectual and developmental disabilities (IDD) and those with limited hearing were significantly less likely to initiate care in the first trimester compared to women without disabilities, and women with IDD had the highest proportion with entry into care delayed until the third trimester ^7^. Although disaggregation by type of disability was beyond the scope of the current study, it is possible that those with IDD were primarily captured in the STAR+ group and were particularly vulnerable to delayed or no receipt of prenatal care.

Our finding that fewer than half of the women received guideline-based preventive screening is lower than the rates observed in linked enrollment, claims, and birth certificate data from Medicaid beneficiaries in Ohio from 2011-2015, where guideline-concordant screening ranged from 50-60% ^31^. This low compliance with recommendations among Medicaid beneficiaries is concerning. TDAP uptake was especially low in our sample, with fewer than one in five women vaccinated during pregnancy. National data show that TDAP vaccination rates during pregnancy have been historically low in the U.S. among both commercial and Medicaid insured patients, increasing from a low of 0.5% in 2010 to 31.4% in 2017 for the latter ^32^.

The policy context in Texas is particularly relevant to our findings. During the study period, pregnant women in Texas qualified for Medicaid at 185–198% of the federal poverty level, whereas nonpregnant adults with disabilities qualified through SSI at approximately 74% of the federal poverty level, allowing continuous enrollment prior to pregnancy ^33^. In theory, this pattern of continuous enrollment should have facilitated more consistent access to care and higher concordance with recommended prenatal screening among STAR+ beneficiaries, as seen in states that expanded Medicaid ^27^. Instead, we observed similar, or in some cases worse, gaps in guideline-concordant care among STAR+ enrollees. This suggests continuous coverage alone may be insufficient to ensure receipt of high-quality prenatal care and points to the presence of structural, clinical, or administrative barriers that persist even among populations with more stable Medicaid coverage. However, income differences between groups may partially explain observed disparities.

Evidence-based obstetric implementation strategies, including audit and feedback, reminders, and other best practices safety bundles, have demonstrated effectiveness in changing clinical behaviors and improving guideline adherence ^34^. For example, maternal quality of care improvement initiatives have had a significant impact in areas such as the timely treatment of hypertensive emergencies during pregnancy and immediately following delivery, and reductions in severe maternal morbidity from obstetric hemorrhage ^35–37^. Our work shows ample opportunity for similar quality improvement approaches for timely prenatal care delivery for high risk groups, such as those with disabilities.

### Limitations

Several limitations warrant consideration. First, the study period (2010–2015) preceded changes such as telehealth expansion and evolving reproductive health policies that may affect access for women with disabilities. Therefore, our findings represent a baseline that may not fully reflect current care patterns, though the structural barriers that hamper high quality care for all low-income women likely persist. An analysis of more recent data will be helpful in understanding trends in prenatal care receipt since 2015. Second, Texas’s decision not to expand Medicaid limits generalizability to expansion states with different coverage pathways and eligibility thresholds. Our results reflect a more restrictive coverage environment and may be most relevant to non-expansion states.

Third, we excluded 376,339 births due to lack of continuous enrollment for three months, including the birth month, representing 33.5% of the initial sample. As shown in Appendix 1, excluded individuals were more often foreign-born Hispanic and had less than a high school education. This likely introduced selection bias toward more stable enrollees and may underestimate barriers to guideline-concordant care. Fourth, some deviations from guidelines may reflect appropriate clinical decision-making not captured in administrative data. Additionally, we could not account for tests ordered but not completed due to patient delays, laboratory errors, or insurance issues, which may differ across disability groups.

## CONCLUSION

The finding that guideline-concordant care was suboptimal in this population underscores systemic quality challenges in Medicaid prenatal care delivery. However, the particularly low receipt of recommended care among women with disabilities, especially those receiving disability benefits, highlights the need for targeted interventions. Healthcare systems must implement systematic quality improvement approaches, enhanced provider training in disability-competent care, and alternative payment models that support comprehensive prenatal care for this vulnerable population. Improving guideline-concordant care delivery represents a critical opportunity to address the documented disparities in maternal and infant outcomes experienced by women with disabilities.

## Data Availability

Data are available upon request from the Texas Department of Health and Human Services.

